# COVID-19. Transport of respiratory droplets in a microclimatologic urban scenario

**DOI:** 10.1101/2020.04.17.20064394

**Authors:** Nicolás Guerrero, José M. Brito, Pablo Cornejo

## Abstract

Although there are some recent studies which intent to address the spread of respiratory droplets through the air, these correspond to indoor conditions or outdoor situations which not take into account realistic scenario. Less attention has been paid to the spread of respiratory droplets in outdoor environments under microclimatologic turbulent wind and which is of growing importance given the current COVID-19 epidemic. We implement a computational model describing a sneezing person in an urban scenario under a medium intensity climatological wind. Turbulence was described with a wall-modeled Large Eddy Simulation model and the spread of respiratory droplets by using a lagrangian approach. Results show the spread of respiratory droplets is characterized by the dynamics of two groups of droplets of different sizes: larger droplets (400 – 900 μm) are spread between 2–5 m during 2.3 s while smaller (100 – 200 μm) droplets are transported a larger range between 8–11 m by the action of the turbulent wind in 14.1 s average. Given the uncertainty of potential contagion over this way and with this reach, these efforts are an intent to contribute to shine a light on the possibility of adopting stricter self-care and distancing measures.

## Introduction

David Heymann, from the London School of Hygiene and Tropical Medicine, who was one of the leaders during the SARS outbreak and public health expert said a few weeks ago that the propagation of COVID-19 in open spaces was one of the important unknowns yet to be realised [1].

The world health organisation (WHO) has communicated recently that for COVID-19 there is no evidence strong enough to adopt measures against the aero-transported contagion. This is, by droplets in the air smaller than 5-10 μm as aerosol, exhaled during breathing or normal speech [2], with exception of close-contact cases, as for care-takers and direct contact with infected persons at a close range (less than 1 meter). Its general recommendations, and thus from several governments and health agencies, is to keep a distance of at least 1 meter from persons who are coughing or sneezing [3]. The discussion about precautionary measures is indeed important, since it might affect the steps taken to order the free or restricted circulation of people in public places and to minimize contagion, including the use of masks, among others. Within the scientific community there is a lack of consensus, and some opinions are controverted. There have been detections of RNA material further apart from the closer radius, and there is a lack of experiments and systematic trials to determine this fact with more precision, for instance, to see if the amount of material found might account for an effective infectious potential [2,4].This takes the discussion further, meaning not only to the scientific community but also to politics and public policy, where precautionary measures can be essential. This study is a consequence of the analysis of scientific literature and news reported by WHO and scientific journals. As a team specialized on the modelling and simulation of fluid flow we could describe and predict some condition related with the spread of respiratory droplets in order to shine a light on one part of the discussion and collaborate to validate or question recommendations to adopt distancing and self-care measures.

The velocity of the exhaled flux of air during sneeze is much higher than in other processes as breathing or even coughing [6,7]. A sneeze exhales more than a million droplets up to a distance of 3 meters. However, the concentration of droplets at 1 meter of distance from its source is reduced down to 0,001% [6], which coincides with the mentioned distancing recommendations of the WHO [2]. Nevertheless, due to Stokes’ drag, the smaller droplets (< 5–10 μm) stay suspended in the air for prolonged periods of time, situation that might eventually conduce to aero-transported transmission. [2,6]. On the other hand, out of 75,465 cases of COVID-19 analysed in China, none of them was attributed directly to this mechanism [2,5]. Specialists from China and the United Kingdom have said that the probability of contracting the virus this way is not completely negligible and that further studies are necessary [4]. The World Health Organisation has emitted an official statement about this situation [2]. However, it has been often misinterpreted by the media. That there is no aero-transported transmission does not mean that the virus transmitted over the air cannot be a carrier of the disease, for instance, in an urban microclimatologic scenario. From the literature and information, we have analysed, this fact has not been explored thoroughly.

## Simulations

Our predictive research was performed using a simulation of the dispersion of polydisperse droplets (multi-dimensional) exhaled during a sneeze in a microclimatologic urban scenario, using a model based on Computational Fluid Dynamics (CFD). Anthropometric variables like exposed oral and nasal area, as airflow, volume fraction and diameter distribution of droplets, and time of duration of sneeze under analysis were taken from the literature [6,7,10]. The microclimatologic scenario related to wind was defined according to [8,9] and they represent a medium intensity including its attenuation respect to the difference between the average velocity magnitude measured in the periphery and the urban nucleus of cities. The turbulence of the microclimatologic wind was simulated using a wall-modeled Large Eddy Simulation. The dispersion of droplets and its interaction with the velocity field was described using a lagrangian approach. The diameter distribution was modeled by a Rosin-Rammler function. These are relevant technical steps towards realistic outdoor conditions.

**Table 1.** Simulated condition: sneeze parameters and site condition.

| Variable | Value | References |
| --- | --- | --- |
| Wind free stream velocity magnitude | 2 [m/s] | [8,9] |
| Free stream turbulence intensity | 2 % | [15] |
| Nasal and oral area | 1,5 y 2,5 [$\text{cm}^2$] | [6] |
| Sneeze total time 1 s | 1 [s] | [6,14] |
| Airflow during sneeze | 250 [lt/min] | [6,14] |
| Volume fraction of droplets | 10e-5 | [10] |
| Diameter distribution of droplets | -- | [7] |

## Results

Our results (see Figure 1 and on-line video1) indicate that the effect of microclimate is very relevant over the propagation of droplets, where dispersion is enhanced by the turbulent wind. In an urban sector under wind of moderate intensity, the respiratory droplets of bigger size (400- 900 μm) and to which the capability of transmitting the disease because of the amount of Virus material they contain, are transported up to a distance of 5 meters. This happens in a short period of time (2.3 s) due to their higher momentum, while the smaller droplets (100-200 μm) can be transported to an even higher distance, reaching an average up to 11 meters in 14.1 s, referential distances are seen in the Figure 1. This is several times over typical precautionary recommendations. In relation to the different analysed contexts, models and measurements performed until now, they include typically indoor conditions or outdoor environments without taking into account the action of turbulent wind [11,12,13]. Although these allow one to get an idea of the dynamics, they are different from a realistic outdoor condition, where human circulation tends to be more unconcerned or unaware with respect to its adverse effects, and might cause a faulty sense of safety.

**Figure 1.**
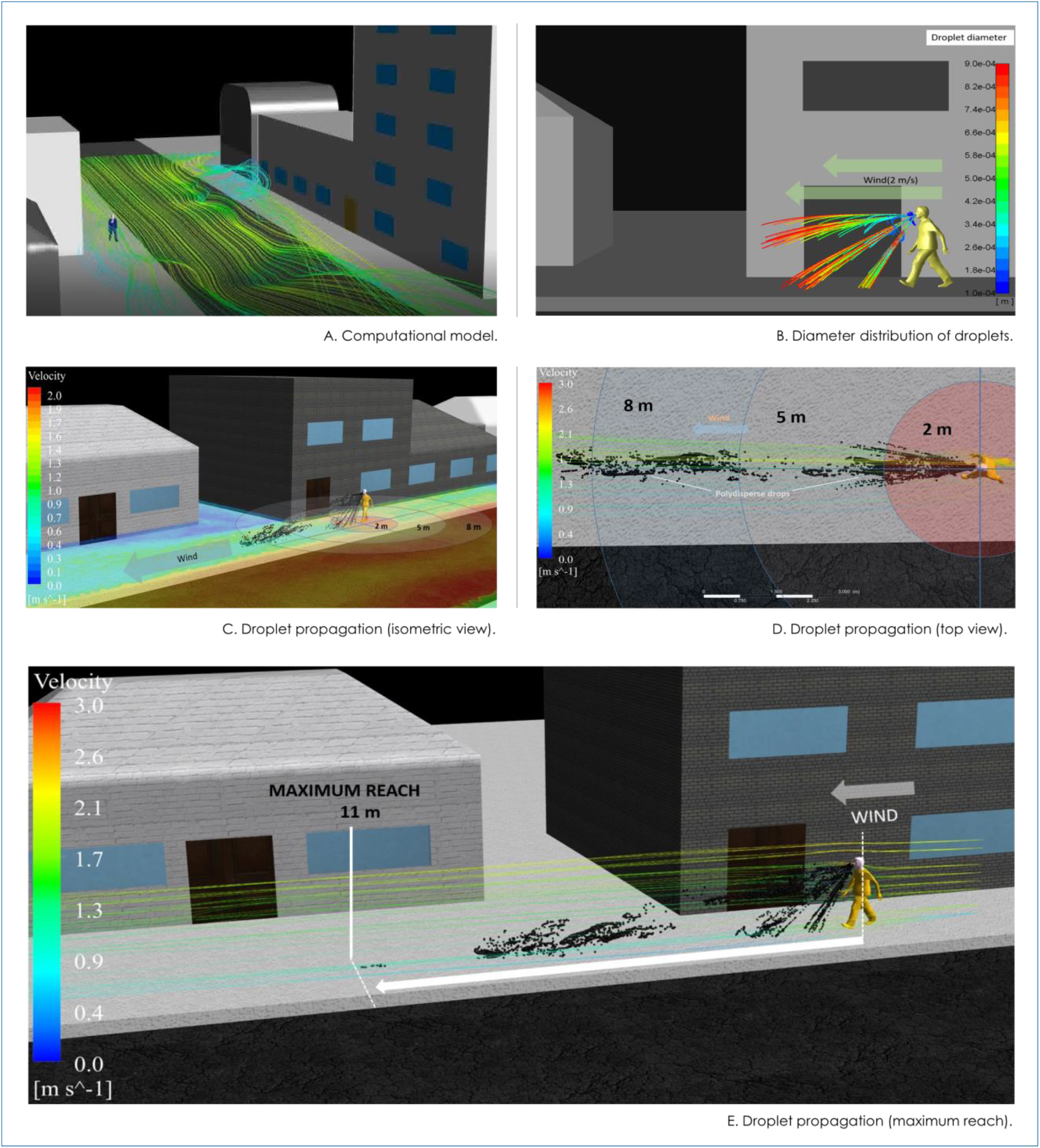
Panel A shows the computational model of the considered urban scenario. The diameter distribution of droplets during the sneeze 0.33 s after model initialization (sneeze total time 1 s) is shown in panel B. Next panels show droplet propagation in the time interval [0.33 - 11.8] s overlapped with contour plot of velocity magnitude of wind (Panel A), pathlines of wind colored by velocity magnitude in top view (Panel D) and pathlines of wind colored by velocity magnitude indicating the maximum reach.

## Conclusions

Our results have allowed us to observe that in different urban zones, under standard microclimatologic conditions, the reach of particles exhaled by a sneezing person and available to be transported by the turbulent wind can be more than 3 times higher than the recommended precautionary distances suggested during the present COVID-19 pandemic. These results represent an everyday urban situation based on the simulated realistic scenario.

Given the uncertainty of potential contagion over this way and with this reach, these efforts are an intent to contribute to shine a light on the possibility of adopting stricter self-care and distancing measures. Nevertheless, in order to determine the real contagion risk related to a certain range of particle diameter at the mentioned distances, infectiology studies have to be performed [3]. This might be difficult because they imply performing tests on living persons or other approaches that are beyond the direct focus of the present study.

## Data Availability

Data available on request from the authors

## Acknowledgments

Author acknowledges Claudio Brito (MSET) for his support on the document edition, Rodrigo Montes (INCAR) for sharing relevant references cited in this short manuscript and Braulio Gatica for his help during the postprocessing and video editing.

## Footnotes

1 https://youtu.be/Rb-xRGPHGPI

## Notes

### Competing Interest Statement

The authors have declared no competing interest.

### Funding Statement

This work was founded by INCAR FONDAP Project N°15110027;CONICYT and MSET Chile SpA.

